# Computer-assisted craniometric evaluation for diagnosis and follow-up of craniofacial asymmetries: SymMetric v. 1.0

**DOI:** 10.1101/19007054

**Authors:** Eduardo Joaquim Lopes Alho, Carlo Rondinoni, Fabio Okuda Furokawa, Bernardo A. Monaco

## Abstract

**Purpose:** The current assessment of patients with craniofacial asymmetries is accomplished by physical examination, anamnesis and radiological imaging.

We propose a semi-automated, computer-assisted craniofacial evaluation (SymMetric v 1.0) based on orthogonal photography of the patient’s head in 3 positions. The system is simple, low-cost, no-radiation or special resources needed. Although it does not substitute CT in cases of doubt between craniosynostosis and positional plagiocephaly, multiple numeric evaluations indicate regional deformities and severity of the asymmetry, which can help in the clinical decision of indicating or not the orthosis in positional deformities, determining treatment duration or evaluating surgical outcomes after correction.

**Methods:** A Matlab-based tool was developed for digital processing of photographs taken in 3 positions (anterior, superior and lateral). The software guides the user to select visible and reproducible landmarks in each photograph acquisition and calculates multiple indexes and metrics, generating a set of comprehensive plots to offer the user an overview of head and facial symmetry across the orthogonal views. For purposes of demonstration, we evaluated 2 patients (one control and one with non-sinostotic deformity).

**Results:** The results show a clear differentiation of the control and plagiocephalic patient metrics mainly in the superior view, showing potential for diagnosis of the condition, and also detected the clinical improvement during helmet treatment in the follow-up, 3 and 5 months after orthosis’ use.

**Conclusion:** We presented a proof-of-concept for a low cost, no radiation evaluation system for craniofacial asymmetries, that can be useful in a clinical context for diagnosis and follow-up of patients.

## 1 Introduction

Craniofacial asymmetries include the premature fusion of one or more cranial sutures (craniosynostosis) and positional asymmetries (positional or deformational plagiocephaly). Treatment depends on the diagnosis, with a surgical approach in craniosynostosis and conservative treatment (cranial orthosis or postural/behavioral changes) in positional deformities. Since the launch of the *Safe to Sleep* campaign (formerly known as *Back to Sleep*) in 1994, aimed to reduce the risk of sudden infant death syndrome (SIDS), the incidence of positional deformities has increased [1–4], raising the parental demand for diagnosis and treatment of this condition [5].

The current assessment of these patients is accomplished by physical examination with anthropometric measurements of the baby’s head using spreading calipers, anamnesis and radiological imaging, as skull computed tomography (CT). Three-dimensional laser head scanning is also available, but associated with higher costs and available only in specialized centers. The most widely used indices used to evaluate a general head shape in a numeric fashion in the clinical setting are the cephalic index (CI), calculated by dividing the maximum cranial width by the length [6] and the cranial vault asymmetry index (CVAI), calculated by the difference in length between two diagonal lines drawn at 30 degrees from the anterior-posterior pole, divided by the shorter of the two diagonals [7]. A systematic review conducted aiming to determine how head shape is measured in positional plagiocephaly [8] concluded that, to identify the appropriate treatment pathways of this condition, categorization correlated to qualitative and quantitative parameters is desirable. The categories should be based on measurements using a reliable, non-invasive and simple measurement technique. Such method is still lacking on the literature.

In this paper, we propose a semi-automated, computer-assisted craniofacial evaluation based on orthogonal photography of the patient’s head in 3 positions. The system is simple, low-cost, no-radiation or special resources needed. The photographs can be easily taken with smartphones in the office or even at home and sent for analysis. Minimal training to obtain the orthogonal views is required. The metric analysis can be performed also in photogrammetry or CT images, although they are not obligatory. We developed multiple numeric evaluations, which indicate locations and severity of the asymmetry and a categorization of severity based on the metrics generated by the software.

## 2 Methods

In this study 2 patients (n=2), one control and one patient with positional plagiocephaly, were submitted to the SymMetric v1.0 evaluation. Control patient (CP) is male, with 4 months old at the evaluation moment, coming to routine pediatric appointment with no complains of skull deformities. A second subject (PP) was referenced from the pediatrician at 4 months of age for evaluation of skull deformity. He had already been submitted to computed tomography (CT), confirming non-synostotic plagiocephaly. Informed consent was obtained for both participants, and photographs in superior, lateral and anterior views were captured. Therapy with helmet was indicated for PP, and new photographs were acquired after 3 and 5 months of helmet therapy. All photographs were purchased with a smartphone in the office, with no additional setup needed. The superior symmetry function was used to compare the control to the PP patient and all three functions were used to compare metrics in the follow-up of helmet treatment. The evaluations were performed after conclusion of the treatment, and did not influence clinical decisions.

### 2.1 Software SymMetric v 1.0

The software was developed in Matlab R_2016b platform. Three functions are executed: superior symmetry, lateral symmetry, and anterior symmetry. Therefore, 4 orthogonal pictures in these views (lateral right and left) are required as input for numeric computations. While opening the GUI (Graphic User Interface), the user is required to choose which functions to process and how many photographs (number of pictures from the same view in different times or different patients) are going to be compared.

#### 2.1.1 Superior Symmetry function (SSF)

This function is the most important while evaluating positional deformities and craniosynostosis. From a superior view (figure 1C), the user is guided to position the midline and contour the baby’s head, in order to segment it from the background. From this input, the following metrics are calculated: cephalic index (CI), cranial vault asymmetry index (CVAI), eccentricity (Ec), superior symmetry index (SSI) and asymmetry severity index (ASI). The first two are classical literature measurements[6, 7, 9] and the remaining are proposed now and will be further explored in the results and discussion sections. This function treats the head’s surface as an ellipsoid, divided in 2 hemispheres by the midline defined by the user. The SSIs are computed by using the centroid distance to the most extreme border of the head on the left side, divided by the distance of the centroid to the most extreme border of the head on the right side in the homologous angle for the contralateral hemisphere (for example 30° and -30°). There are 180 indices, comparing each degree from the left hemisphere to the homologous degree in the right hemisphere. In a perfect symmetric ellipse, all the SSIs are equal to 1. In our case, it is assumed that the maximum difference between the ratio of two homologous diagonals should not exceed 3.5% [7] in symmetric heads. Therefore, our normality values are expected to be in the 0.965 - 1.035 range. Using the homologous diagonals at 30°, the CVAI is also calculated. In order to evaluate severity, an Asymmetry Severity Index (ASI) is calculated, by the sum of all SSIs found outside the normality values. We classified 4 zones, being 0%-5% considered normal, between 6%-25% focal asymmetry, 26%-50% intermediate asymmetry and above 50%, hemispheric asymmetry. Plots for all these metrics are displayed automatically.

**Fig. 1.**
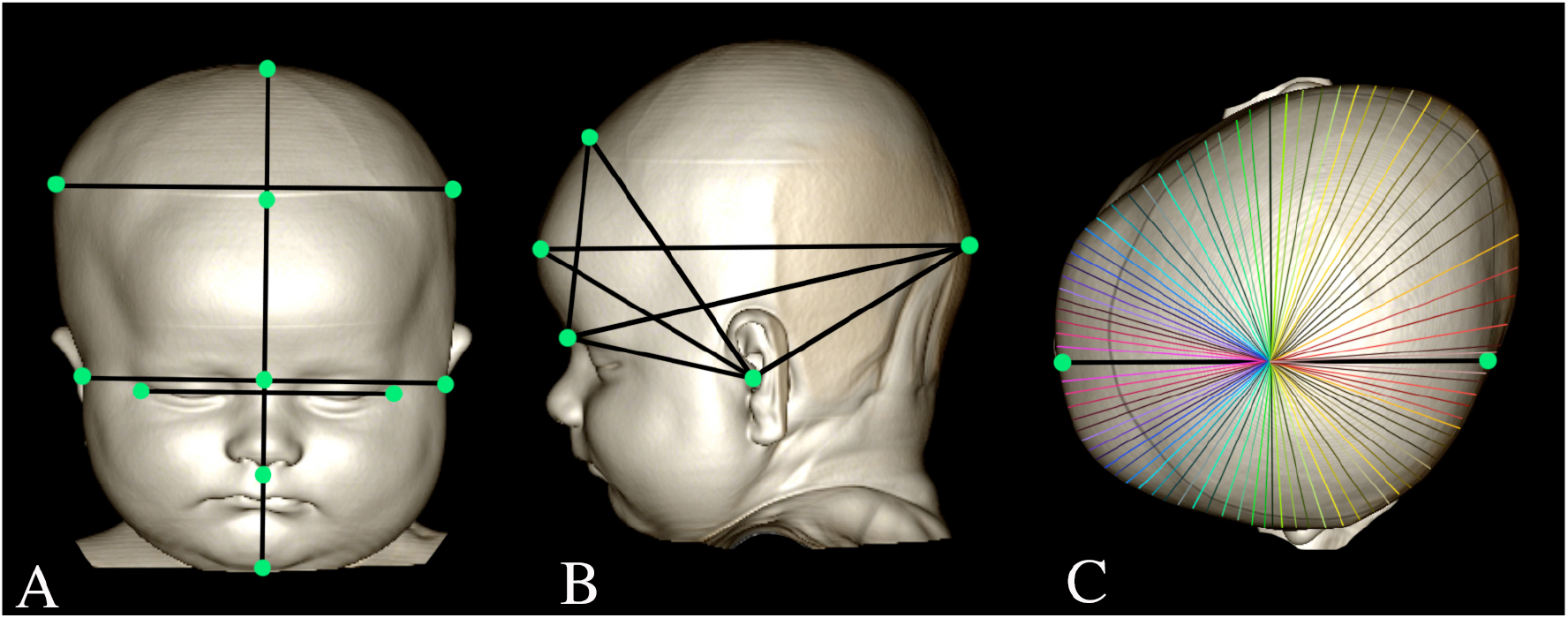
A) Anterior view used for ASF and facial landmarks (green) and distances (black) computed B) lateral view used for LSF C) Superior view used for SSF. Two points in the midline are chosen by the user, and homologous distances (here displayed with the same colors for right and left hemisphere) are compared in the 360°

#### 2.1.2 Lateral symmetry function (LSF)

This function aims to evaluate the craniofacial proportions from a lateral perspective. The user is also guided to select visible landmarks in the lateral view, that will be used to calculate the proportions (figure 1B). The calculated values are summed into an index called Global Head Growth (GHG), in a way to normalize these distances in relationship to a global change of the skull due to normal head growth in time. By dividing each of those distances by the GHG and multiplying by 100, an index of the contribution of each distance to the head proportions (figure 1 B) is obtained.

#### 2.1.3. Anterior symmetry function (ASF)

In the same way as the previous functions, the user is required to set visible landmarks such as glabella, sub nasal point, biparietal points, tragus and epicantus (figure 1A).

The proportions between face’s height and width (facial ratio) and eye distance and facial width (eye ratio) are computed. In proportional faces, those ratios are close to 1.618 (fi or Golden ratio)[10]. The face is also divided in thirds, such as an ideal proportion is a 1:1:1 ratio [11]. The proportions between biparietal distance and facial width are also calculated, ideally slightly greater than 1 (craniofacial proportion). Finally, the angles between eyes and ears in relationship to midline are also computed, and should be approximately 90*°*. Very different values indicate asymmetric ear implantation or orbital/ cranial deformations with orbital impact.

All these indices and proportions should be evaluated evolutionarily in the same patient, to assess effectiveness of treatments.

## 3 Results

### 3.1 Positional plagiocephaly x control evaluation

For this comparison, the SSF was applied to PP at diagnosis and CP at 4 months of age. The cephalic index (CI) calculated for CP was 78,97% and for PP 87,76% (figure 2A). The normality range is between 75%-85%[12]. The CI for CP was considered to be in the normal range, but for PP was higher, indicating brachycephaly. Eccentricity of a conic section is a measurement that characterizes its shape. The eccentricity of a circle is zero and of an ellipse is greater than zero and less than one. The conic section of a skull can be considered as an ellipse and we use this metric to evaluate head elongation. In brachycephalic heads, the values tend to be closer to 0 and in dolicholcephalic heads, closer to 1. For our subjects, the measured Ec for CP was 0,60 and 0,47 for PP. These findings confirm that PP’s head shape is less elongated (brachycephalic) than CP’s. The SSIs showed a clear difference between CP and PP (figure 2B). While all 180 SSIs for CP were in the normality range (0.965-1,035), for PP, the values considered to be normal were just the one closer to midline, rendering a sinusoid curve with greater discrepancies in the left frontal area (between 30° to 60°) and right parietal (120° to 160°) area, with values reaching 0,90 and 1,13. The CVAI was calculated as described in [7], being in the normality range for CP (0.20%), and above the 3.5% asymmetry threshold for PP, with an index of 8,29%.

**Fig. 2.**
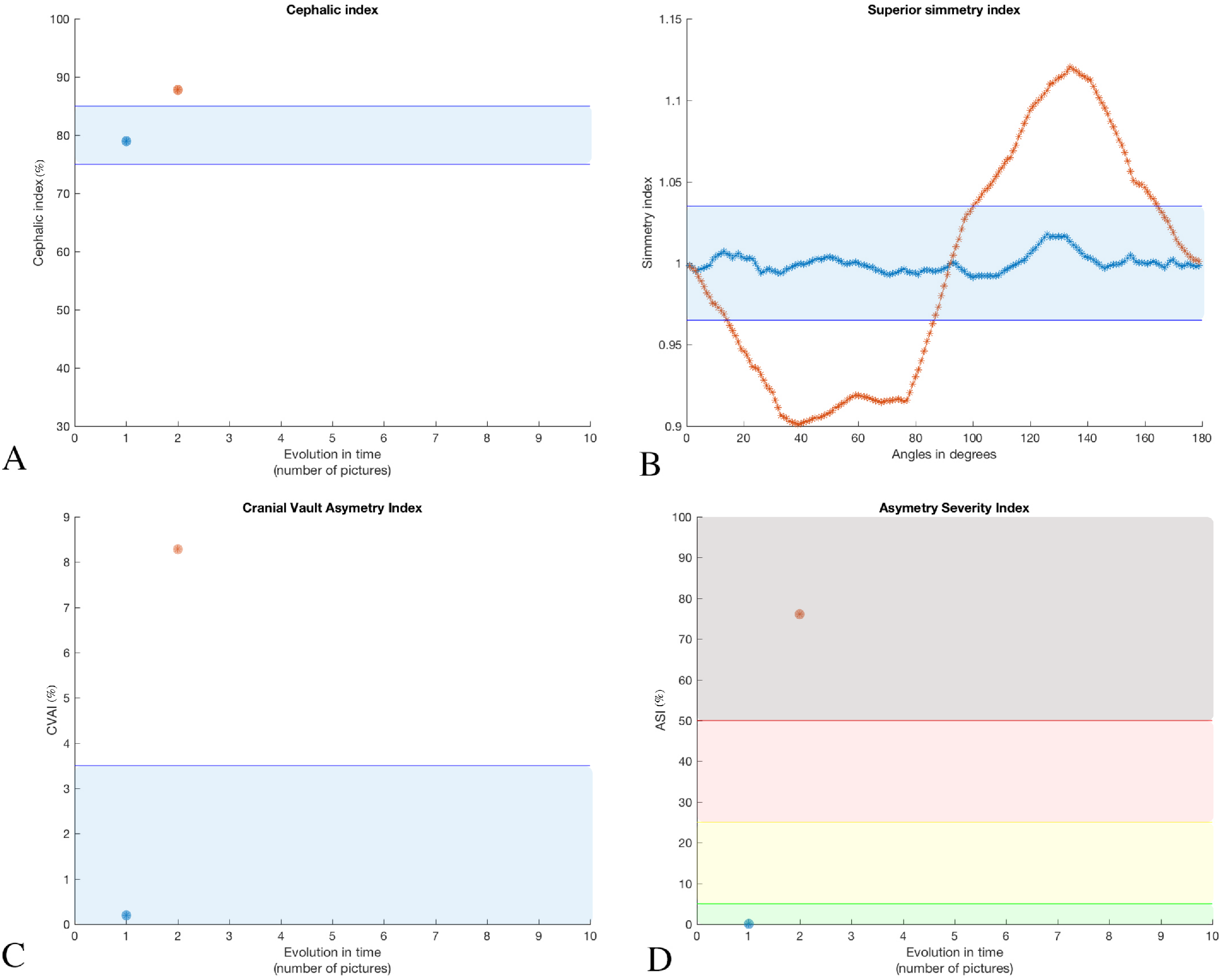
Metrics for control patient (CP) represented by blue circles and asterisks, and plagiocephalic patient (PP), represented by the orange circles and asterisks A) Cephalic index for CP is within the normality range (75%-85%), while PP is above B) Superior symmetry indices comparing 180° of each hemisphere. CP is within the blue zone (normality range) while PP has a sinusoid curve outside normal boundaries C) CVAI for CP inside the blue zone, while PP is above the limit D) Asymmetry Severity Index with categorization divided in normal (green zone), focal asymmetry (yellow zone), intermediate asymmetry (red zone) and hemispheric asymmetry (brown zone) Note CP is in the green zone, while PP in brown zone

The Asymmetry Severity Index (ASI) was calculated for both subjects, with 0,00% for CP (no asymmetry) and 76,11% for PP, in the hemispheric asymmetry zone (more than 50% of the SSIs outside the normal range) (figure 2D).

### 3.2 Evolutional metrics of positional plagiocephaly during helmet treatment

The objective of this analysis is not to evaluate helmet treatment for PP, but instead, to apply the metrics proposed in this study to verify if they would reflect the clinical outcome observed. Treatment was carried out for 6 months, 23h a day. Parents and pediatrician were satisfied with the aesthetic final result, and the deformity was very discrete with 10 months of age. Photographs were taken at diagnosis and 1 month after (but still no helmet), 2 months and 5 months after orthosis use.

The evolutional analysis of the SSIs showed a progressive flattening of the sinusoid curve obtained at the moment of diagnosis, reaching the normality range for the majority of values. The maximum SSI value dropped from 1,170 on the first evaluation to 1,050 (upper limit 1,035) on the final evaluation and the minimum value raised from 0,870 to 0,950 (lower limit 0,965) after 5 months of therapy (figure 3A). The CVAIs also dropped from 9,85% to 5,20% (3 months) and 2.99% (5 months), entering in the normality range (<3.5%) (figure 3B). The ASI also dropped dramatically from 78,90% (diagnosis) to 64,40% (3 months) and 39,40% (5 months) with helmet use, with reclassification of PP from hemispheric asymmetry to intermediate asymmetry (figure 3C). Interestingly, the cephalic index remained above the superior threshold even in the third evaluation (figure 3D).

**Fig. 3.**
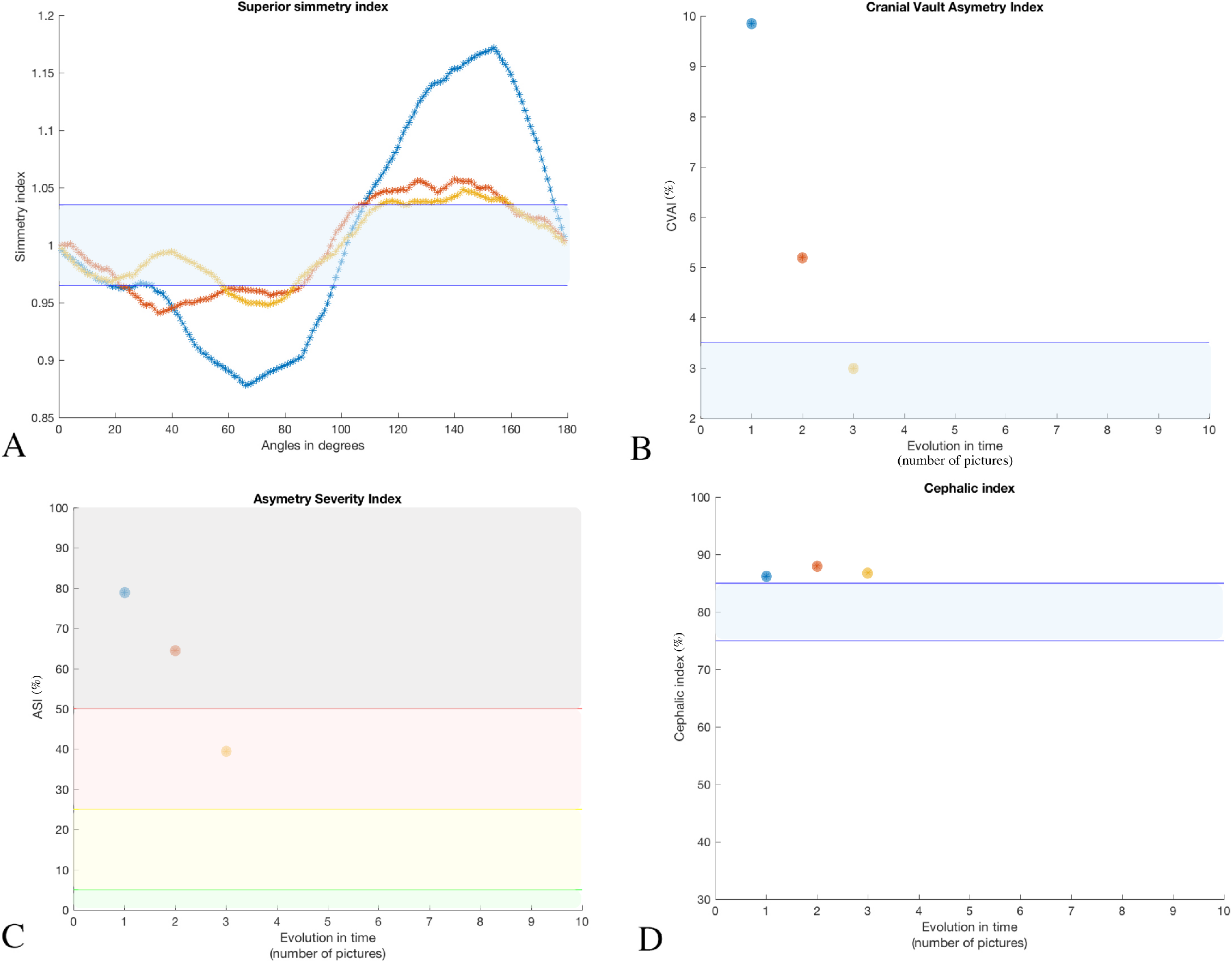
Metrics at zero-time (1- blue asterisks), 3 months (2- orange asterisks) and 5 months (3- yellow asterisks) follow-up A) Results of SSIs for PP), showing flattening of the initial sinusoid curve, almost fitting entirely in the normality range (blue zone) B) CVAI, entering into the normality zone (blue) at time 3 C) ASI leading to reclassification of the severity category at time 3 D) Cephalic index was maintained above the normality range (blue zone) even after effective treatment

The craniofacial proportions (biparietal/facial width ratio) calculated by the ASF were slightly greater than 1 in all measurements, showing proportional head size to the facial width (figure 4A). Regarding facial proportions divided in thirds, in the first evaluation the proportions were close to 1:1:1. At 3 months after helmet use those proportions changed slightly, with the superior third more prominent than the inferior third. On the last measurement, those proportions had a tendency of returning to baseline (figure 4B). A possible explanation for that would be physiological growth of the neurocranium on the first months of life. This hypothesis has to be verified by analyzing several subjects over the first year of life.

**Fig. 4.**
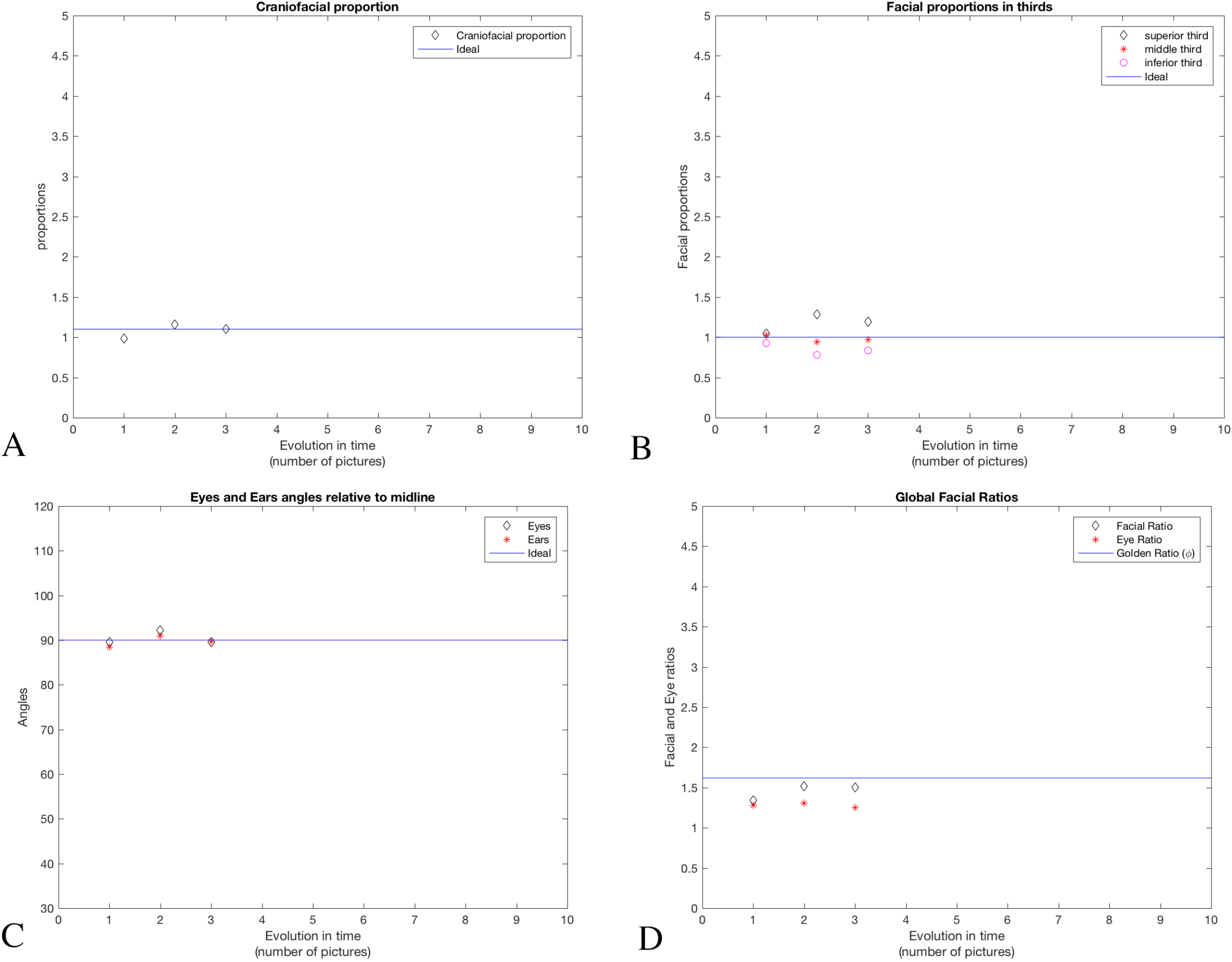
Metrics at zero-time (1), 3 months (2) and 5 months (3) follow-up. Craniofacial proportions are not severely altered in PP patient. Small evolutionary differences are discussed in text

The angles of eyes and ears relative to midline were close to 90° in all three evaluations, showing no orbital or ear implantation issues due to plagiocephaly (figure 4C). The facial (facial width/facial length) and eye (distance between eyes/facial width) ratios were stable, with values close to the Golden ratio in all measurements (figure 4D).

Finally, the metrics generated by the LSF showed as main change the raise of the contribution of the meato-bregmatic and nasio-bregmatic distances, with decrease of the nasio-meatal contribution to global head growth (figure 5). These findings indicate growth of the anterior parts of the neurocranium, without raise in facial contribution, supporting the hypothesis that in the first months of life, the neurocranium must grow in a faster rate as the viscerocranium in order to accommodate a fast-growing brain.

**Fig. 5.**
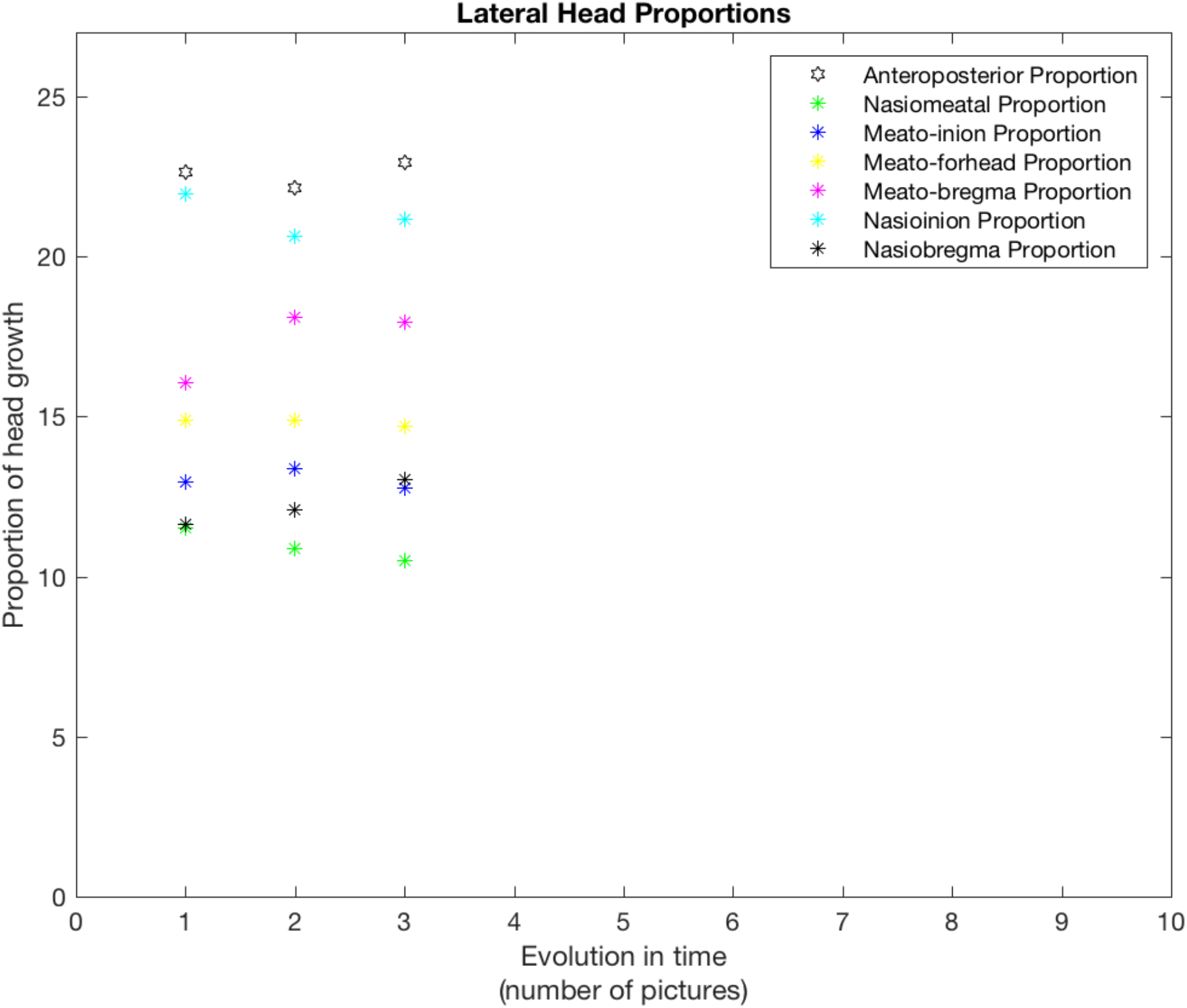
Lateral Head proportions at zero-time (1), 3 months (2) and 5 months (3) follow-up. Several distances are computed proportionally to global head growth. Some proportions changed over time, possibly indicating physiological growth of the neurocranium to accommodate brain growth.

## 4 Discussion

In this preliminary conceptual study, we developed a software that aims to produce metrics for categorization of cranial asymmetries, with several quantitative parameters, that can be helpful for decision making in the clinical environment. A simple and non-invasive technique was used to analyze two individuals, and the metrics obtained allowed to differentiate a control from a positional plagiocephalic patient, categorize severity of deformation, and characterize it in a consistent fashion. A clear metric modification in the follow-up treatment was also observed. It is a comprehensive tool, since different functions analyze different aspects of head shape, being able to distinguish among several conditions.

To date, the available methods for diagnosis and follow-up of those patients are performed either in specialized centers equipped with 3D laser scanners or 3D photogrammetric studios, or simply by clinical examination calculating CI and CVAI and subjective criteria. The parameters defined in our study are complimentary to the current indices, and can enhance the precision of diagnosis and follow-ups after either conservative or surgical treatment. Meanwhile the CVAI evaluates the symmetry at 30° diagonals, the SSIs evaluate the head in its 360°, detecting focal asymmetries that might not alter significantly the diagonals at 30°. A good example of the importance of more parameters is that while during helmet treatment the cephalic index in the PP patient was stable above normal limits (figure 3D), all the other metrics showed massive changes (figure 3A, B and C), in agreement with the clinical impression. Therefore, the CI might not be a reliable metric for follow up in some cases. In the PP patient it was possible to observe progression of several metrics and some variations of the proportions in the lateral and anterior views could be explained by physiological head growth, but this hypothesis can only be verified after validation of those parameters in a normal pediatric population, building normality curves for each of those indices.

There are some limitations, however, of SymMetric v 1.0, that will be overcome in the next versions. The need of definition of landmarks by the user and obtaining orthogonal pictures in the same positions require some training and practice. Great variability in choosing landmarks and obtaining images by a non-trained user can impact on final results. Therefore, for version 2.0, automatic recognition of the landmarks will be performed by implementation of a Scale-Invariant Feature Transform (SIFT) algorithm [13]. This algorithm allows recognition of points regardless rotation, translation, scaling and are partially invariant to illumination changes and affine or 3D projection. These characteristics make it suitable for facial recognition[14] in such context. The implementation of a colored cap with fixed fiducials can also contribute to ameliorate automatic skull segmentation (color-based k-means clustering) and metrics computation. Versions for desktop and mobile phones can be developed to facilitate access of a broader number of professionals. A database can be built in a cloud compliance environment and used to train artificial neural networks designed to enhance the precision of detection of points, diagnosis and follow-up.

## Data Availability

Data are available upon reasonable request

## 5 Conclusion

SymMetric v 1.0 is a comprehensive tool for semi-automated craniometric evaluation of patients with craniofacial asymmetries. Its application was illustrated with 2 subjects, showing clear differentiation of a control patient from non-sinostotic plagiocephaly patient, and revealed sensitive metric changes in the course of treatment. It can be a helpful tool for clinicians in their daily practice.

